# Integrated Right-Heart Remodeling Phenotypes and Prognosis in Tricuspid Regurgitation: An Automated Strain Echocardiography Study

**DOI:** 10.64898/2026.05.28.26354377

**Authors:** Jiesuck Park, Soongu Kwak, Yeonyee E. Yoon, Jun-Bean Park, Jiyeon Kim, Jaeik Jeon, Yeonggul Jang, Seung-Ah Lee, MinJung Bak, Hong-Mi Choi, In-Chang Hwang, Seung-Pyo Lee, Hyung-Kwan Kim, Yong-Jin Kim, Goo-Yeong Cho

## Abstract

**Background:** Echocardiographic assessment of tricuspid regurgitation (TR) remains valve-centric, and right-heart remodeling is not captured. Strain parameters carry prognostic value but are evaluated in isolation.

**Objectives:** To develop integrated right atrial (RA) and right ventricular (RV) remodeling indices using automated echocardiography and assess their utility for TR severity grading, phenotyping, and prognostic stratification.

**Methods:** We analyzed 8,231 patients with functional TR (mild-or-greater) from two tertiary centers (2023–2024) using an automated AI-based echocardiographic solution. The RA remodeling index (RA reservoir strain/RA volume index) and RV remodeling index (RV free wall strain/RV end-diastolic area) were derived automatically; patients were classified into four RA–RV remodeling phenotypes. The primary outcome was all-cause death or heart failure (HF) hospitalization.

**Results:** During median follow-up of 19.3 months, the primary outcome occurred in 574 patients (7.0%). Both indices outperformed individual components for severe TR discrimination (RA: AUC 0.857 vs. 0.757; RV: 0.710 vs. 0.601; both *P*<0.05). After multivariate adjustment, the RA (HR per unit decrease, 1.27; 95% CI, 1.09–1.49; *P*=0.002) and RV remodeling indices (2.32; 1.76–3.06; *P*<0.001) were independently associated with the primary outcome; on mutual adjustment, only the RV index retained significance and provided incremental prognostic value (ΔC-index +0.010; NRI +0.237; both *P*<0.05). The four phenotypes showed progressively divergent risk (log-rank *P*<0.001), with combined remodeling (Low RA/Low RV) carrying the highest risk.

**Conclusions:** Automated integrated RA and RV remodeling indices improved TR severity discrimination and enabled clinically meaningful right-heart phenotyping. The RV index conferred incremental prognostic value, whereas the RA index better reflected atrial-stage remodeling and disease burden.

**CONDENSED ABSTRACT:** In 8,231 patients with functional tricuspid regurgitation from two tertiary centers, we derived integrated right atrial (RA) and right ventricular (RV) remodeling indices combining strain with chamber structure using automated echocardiography. These indices outperformed individual strain parameters for severe TR discrimination and stratified patients into four prognostically distinct RA and RV remodeling phenotypes. Both indices independently predicted the composite of all-cause death and heart failure hospitalization. The RV index emerged as the dominant prognostic marker with incremental value over the clinical model, whereas the RA index primarily characterized atrial-stage remodeling and disease burden.

## INTRODUCTION

Tricuspid regurgitation (TR) has historically been regarded as a benign secondary finding, the “forgotten” valve disease, accorded limited clinical attention compared with left-sided valvular pathology.(1,2) Accumulating evidence, however, has established TR as an independent predictor of heart failure (HF) hospitalization and all-cause mortality, with a rising prevalence in an aging population,(3–5) necessitating proactive evaluation and management.

Transthoracic echocardiography (TTE) is the primary imaging modality for TR assessment, providing Doppler-based quantification for severity grading and etiologic classification.(6) However, these frameworks have evolved predominantly in a valve-centric manner, and the structural remodeling and functional reserve of the right-heart system associated with TR have not been sufficiently addressed. The right atrium (RA) and right ventricle (RV) undergo progressive, mechanistically linked remodeling in TR, encompassing RA dilation with loss of atrial compliance, RV enlargement, and longitudinal dysfunction. However, these changes are not systematically captured by current grading schemes.(7,8)

Speckle-tracking–derived strain parameters have demonstrated clinical utility in quantifying right-heart remodeling beyond conventional indices. RV free wall strain (RVFWS) is independently associated with adverse outcomes in TR, reflecting RV longitudinal dysfunction.(9,10) RA reservoir strain (RASr), a measure of atrial capacitance during ventricular systole, extends beyond volumetric assessment by capturing atrial myopathy, and carries independent prognostic value in TR.(11,12) Despite these advances, RA and RV parameters have largely been evaluated in isolation, and a comprehensive framework for integrated right-heart remodeling assessment in TR has not been established. Furthermore, the manual measurement of these parameters remains labor-intensive and subject to inter-observer variability, which may limit their systematic application in routine clinical practice. Recent advancements in artificial intelligence (AI) have enabled automated and reproducible quantification of cardiac structure and function from echocardiographic images, ranging from valvular heart diseases to cardiomyopathies, offering a potential solution to these practical constraints.(13–18)

In the present study, we aimed to develop integrated RA and RV remodeling indices combining strain-derived functional parameters with structural measures, using automated echocardiographic analysis in a large real-world multicenter cohort. Specifically, this study focused on: (1) characterizing RA and RV remodeling across TR severity and defining clinically distinct RA–RV remodeling phenotypes using these integrated indices; and (2) evaluating their independent and additive prognostic value for clinical outcomes.

## METHODS

### Study Population

We retrospectively analyzed consecutive patients from two tertiary medical centers, Seoul National University Hospital (SNUH) and Seoul National University Bundang Hospital (SNUBH), who underwent TTE between January 2023 and December 2024 and were found to have TR of any severity. When multiple TTE examinations were available, the earliest one was selected as the index study for each patient. Of 23,407 identified patients, those with previous tricuspid valve (TV) surgery or intervention (N=184), non-functional TR etiology such as TV prolapse, infective endocarditis, and rheumatic disease (N=50), less-than-mild TR (N=12,593), cardiac implantable electronic device–related TR (N=529), significant left-sided valvular heart disease (N=1,428), or unavailable clinical data (N=69) were excluded, yielding 8,554 eligible cases (**Figure 1**). The study protocol was approved by the Institutional Review Board of each participating institution (SNUH H-2506-143-1652 and SNUBH B-2605-1047-405), with a waiver of informed consent granted due to the retrospective study design.

**Figure 1.**
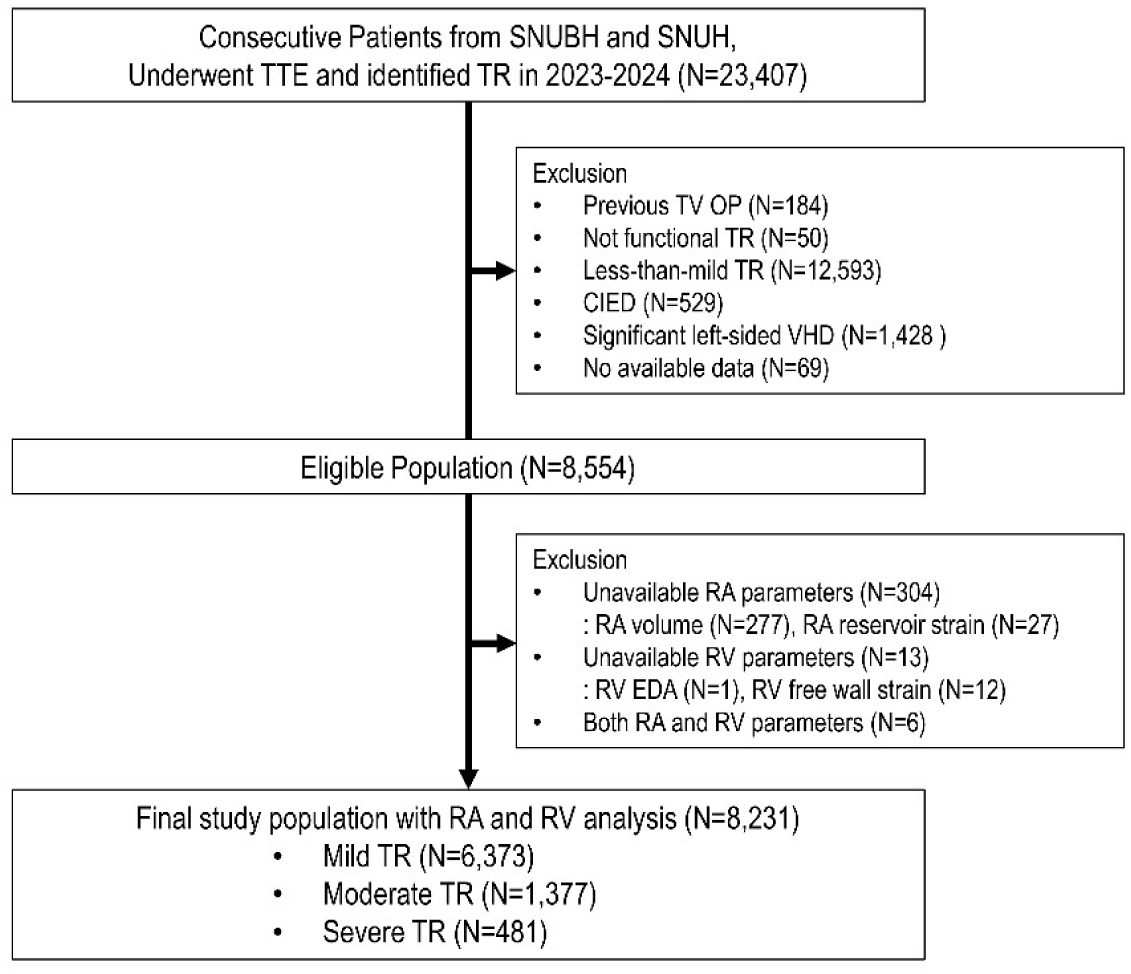
Study Flow. Consecutive patients from SNUH and SNUBH who underwent TTE between January 2023 and December 2024 were screened for TR. Stepwise exclusion criteria are depicted, resulting in 8,554 eligible cases. Among these, patients with unavailable RA or RV parameters due to insufficient image quality were further excluded, yielding a final study population of 8,231 patients. SNUH = Seoul National University Hospital; SNUBH = Seoul National University Bundang Hospital; TTE = transthoracic echocardiography; TR = tricuspid regurgitation; CIED = cardiac implantable electronic device; VHD = valvular heart disease; RA = right atrium; RV = right ventricle.

### Echocardiographic Assessment

All echocardiographic examinations were performed by trained echocardiographers or cardiologists and interpreted by board-certified cardiologists specialized in echocardiography, in accordance with current guidelines as part of routine clinical care.(19) TR severity was evaluated by expert assessment using a multiparametric approach in accordance with current recommendations.(6–8) Qualitative evaluation based on color flow jet characteristics served as the primary basis for TR assessment. When technically feasible and clinically indicated, semiquantitative parameters (vena contracta width) and quantitative parameters (effective regurgitant orifice area [EROA] and regurgitant volume, derived using the proximal isovelocity surface area method) were additionally obtained. Final TR severity was graded as mild, moderate, or severe based on integrated judgment across all available parameters. When quantitative assessment was performed, moderate TR was defined as EROA 0.20–0.39 cm^2^ or regurgitant volume 30–44 mL, and severe TR as EROA ≥0.40 cm^2^ or regurgitant volume ≥45 mL.

### Automated Measurement of RA and RV Parameters

All key RA and RV parameters, including RVFWS, RV end-diastolic area (RVEDA), RASr, and RA volume, were measured using a previously developed AI-based automated TTE analysis system (Sonix Health; Ontact Health Co., Ltd., Seoul, Republic of Korea) that automatically performs view classification, chamber segmentation, image quality (IQ) assessment, and parameter derivation without human interaction.(16–18) The core pipeline has been validated across cardiovascular conditions.(13–18) For the present study, the same framework was applied to derive RASr and RA volume index (RAVI) as research-specific parameters. Pipeline operation, view selection, and IQ-based exclusion criteria are detailed in the **Supplementary Methods**.

RVFWS was defined as systolic deformation of the RV free wall segments across the cardiac cycle, expressed as an absolute value. RVEDA was the RV chamber area at end-diastole. RASr was defined as deformation from end-diastole (smallest RA volume) to peak RA size, capturing reservoir function. RA volume was calculated at end-systole using single-plane Simpson’s method on the RA segmentation boundary and indexed to body surface area to yield RAVI.

Applying the automated quality criteria, 323 of the 8,554 eligible patients were excluded due to insufficient IQ (304 with unavailable RA parameters, 13 with unavailable RV parameters, 6 with both). The final population comprised 8,231 patients (96.2% of eligible; **Figure 1**). All automated measurements were generated before outcome modeling, with clinical outcome analyses performed independently by the academic investigators.

### Construction of Integrated Remodeling Indices

TR-related remodeling involves both structural enlargement and functional deterioration of RA and RV, which are mechanistically linked but do not progress uniformly.(9–12) Strain-derived or volumetric parameters individually capture only one dimension. Ratio-based indices integrating function and structure have been proposed (e.g., RA stiffness index, RV strain-to-pulmonary pressure ratio), but have been evaluated in isolation and do not provide an integrated RA–RV framework.(20,21)

To address this gap, we constructed integrated RA and RV remodeling indices by normalizing each strain-derived functional parameter to its corresponding structural measure. The RA remodeling index was defined as the ratio of RASr to RAVI, reflecting RA functional capacity relative to the degree of atrial dilation. The RV remodeling index was defined as the ratio of RVFWS to RVEDA, reflecting RV longitudinal functional capacity relative to the degree of ventricular enlargement, consistent with current recommendations for RV structural assessment.(19) Higher values indicate preserved functional reserve relative to structural remodeling, whereas lower values reflect disproportionate functional impairment.

Patients were subsequently classified into four RA–RV remodeling phenotypes based on the median values of the RA and RV remodeling indices as cutoff thresholds: the Preserved phenotype (High RA / High RV), the RA-predominant remodeling phenotype (Low RA / High RV), the RV-predominant remodeling phenotype (High RA / Low RV), and the Combined remodeling phenotype (Low RA / Low RV). The Preserved phenotype represents the most preserved right-heart functional state, whereas the Combined remodeling phenotype reflects the most advanced concurrent atrial and ventricular remodeling.

### Clinical Outcomes

The primary outcome was a composite of all-cause death and hospitalization for HF, with each component evaluated individually as a secondary outcome. HF hospitalization was defined as an admission due to HF aggravation with clinical evidence of cardiac decompensation. Clinical events were ascertained through dedicated electronic medical record review. Follow-up duration was calculated from the date of index TTE to the date of the first event or last follow-up, whichever came first.

### Statistical Analysis

Clinical data are presented as median with interquartile range (IQR) for continuous variables and frequency (percentage) for categorical variables. Differences across TR severity groups were assessed using the Kruskal-Wallis test for continuous variables and the chi-square test for categorical variables. Trends in RA and RV strain, structural parameters, and remodeling indices across TR severity were visualized using violin plots; the correlation between RA and RV remodeling indices was depicted as a two-dimensional remodeling map.

Discriminative performance for TR severity was assessed using AUC for severe versus non-severe TR and moderate-or-greater versus mild TR, with pairwise comparisons against individual strain parameters performed using the DeLong method. Ordinal c-statistics across the three-level classification (mild/moderate/severe) were additionally calculated.

Baseline characteristics across the four RA–RV remodeling phenotypes were compared and visually summarized as a heatmap of z-score standardized values to highlight distinctive clinical profiles.

Kaplan-Meier estimates were compared by log-rank test across TR severity, median-dichotomized indices, and the four phenotypes (Preserved as reference). Cox proportional hazards regression assessed associations with the primary outcome, adjusted for age, sex, AF, LV ejection fraction, and TR grade; HRs (95% CI) are reported per 1-unit decrease, with per-1-SD HRs in supplementary analyses. Proportional hazards was verified by Schoenfeld residuals. The same adjustment was used for secondary outcomes (all-cause death, HF hospitalization), with Fine-Gray models applied to HF hospitalization (all-cause death as competing event). Sensitivity analyses additionally treated TV surgery as a competing event for the primary and both secondary outcomes.

Incremental prognostic value beyond the reference clinical model was assessed by changes in c-statistics, time-dependent AUC, and category-free net reclassification improvement (NRI). Pre-specified subgroup analyses (TR severity, AF status, pulmonary hypertension, diuretic use) were performed with interaction tests. Center-related sensitivity analyses comprised discriminative performance by institution, center-stratified event rates, and a center-adjusted Cox model.

All analyses were performed using Python (version 3.12.3). Two-sided *P*<0.05 was considered statistically significant.

## RESULTS

### Study Population

The final cohort comprised 8,231 patients with functional TR (mild, N=6,373; moderate, N=1,377; severe, N=481). Median follow-up was 19.3 months (IQR, 11.2–26.2). Baseline characteristics across TR severity are summarized in **Table 1**. The median age was 72.0 years (IQR, 64.0–80.0), 44.7% were female, and AF prevalence rose with severity (31.9%, 56.1%, 76.9% in mild, moderate, severe TR; *P*<0.001). Left ventricular (LV) ejection fraction, left atrial (LA) volume index, E/e’, TR velocity, and RV systolic pressure all worsened progressively with TR severity (all *P*<0.001).

**Table 1.** Baseline Clinical and Echocardiographic Characteristics According to TR Severity.

| **Variable** | **Mild TR**<br>**(N=6,373)** | **Moderate TR**<br>**(N=1,377)** | **Severe TR**<br>**(N=481)** | **P** |
| --- | --- | --- | --- | --- |
| **Clinical Variables** |  |  |  |  |
| Age, years | 72.0 (64.0–80.0) | 75.0 (67.0–82.0) | 78.0 (70.0–83.0) | <0.001 |
| Female | 2,871 (45.0) | 594 (43.1) | 216 (44.9) | 0.431 |
| Body mass index, kg/m<sup>2</sup> | 23.3 (21.2–25.6) | 23.3 (21.0–25.4) | 23.5 (21.3–25.7) | 0.249 |
| Hypertension | 2,077 (32.6) | 542 (39.4) | 187 (38.9) | <0.001 |
| Diabetes mellitus | 1,101 (17.3) | 285 (20.7) | 91 (18.9) | 0.009 |
| Atrial fibrillation | 2,035 (31.9) | 773 (56.1) | 370 (76.9) | <0.001 |
| **Echocardiographic Parameters** |  |  |  |  |
| LV ejection fraction, % | 60.4 (56.0–64.6) | 59.2 (55.3–63.5) | 59.0 (53.9–63.4) | <0.001 |
| LA volume index, mL/m<sup>2</sup> | 41.4 (31.9–56.8) | 55.0 (39.6–75.7) | 69.5 (53.8–93.8) | <0.001 |
| E/e' | 10.9 (8.5–14.8) | 12.0 (9.0–16.6) | 12.1 (9.5–17.7) | <0.001 |
| TR velocity, m/s | 2.50 (2.30–2.80) | 2.80 (2.50–3.10) | 2.80 (2.50–3.20) | <0.001 |
| RV systolic pressure, mmHg | 32.0 (28.0–39.0) | 38.6 (32.0–47.0) | 41.4 (35.0–51.0) | <0.001 |
| **Auto-measured RA and RV indices** |  |  |  |  |
| RV end-diastolic area, cm<sup>2</sup> | 19.9 (16.4–23.8) | 21.2 (17.4–26.2) | 25.3 (19.9–32.1) | <0.001 |
| RA volume index, mL/m<sup>2</sup> | 25.1 (18.7–34.7) | 37.8 (26.5–55.0) | 64.8 (45.8–95.9) | <0.001 |
| RV free wall strain, % | 17.7 (13.2–21.9) | 16.9 (12.3–21.0) | 15.0 (10.2–19.4) | <0.001 |
| RA reservoir strain, % | 25.0 (15.1–34.2) | 16.1 (10.4–28.1) | 11.8 (8.3–16.7) | <0.001 |
| **Integrated Remodeling Indices** |  |  |  |  |
| RA remodeling index | 1.05 (0.48–1.64) | 0.42 (0.20–0.94) | 0.17 (0.10–0.30) | <0.001 |
| RV remodeling index | 0.87 (0.63–1.12) | 0.76 (0.52–1.00) | 0.57 (0.38–0.78) | <0.001 |

Data are presented as the median (interquartile range) for continuous variables and number (percentage) for categorical variables.
Abbreviations: LA, left atrium; LV, left ventricle; RA, right atrium; RV, right ventricle; TR, tricuspid regurgitation.

### RA and RV Remodeling Indices Across TR Severity

Distributions of individual RA and RV parameters and integrated remodeling indices are shown in **Figure S1**. The RA remodeling index had a median of 0.87 (IQR 0.34–1.51) and the RV remodeling index a median of 0.84 (IQR 0.59–1.09). Both indices showed graded worsening with increasing TR severity (**Table 1**, **Figure 2**): RASr decreased and RAVI increased progressively from mild to severe TR (both *P* for trend <0.001), yielding a marked decline in the RA remodeling index (*P* for trend <0.001); similarly, RVFWS decreased and RVEDA increased (both *P* for trend <0.001), with a corresponding decline in the RV remodeling index (*P* for trend <0.001). The two-dimensional remodeling map (**Figure S2**) shows a progressive shift toward lower index values with increasing TR severity.

**Figure 2.**
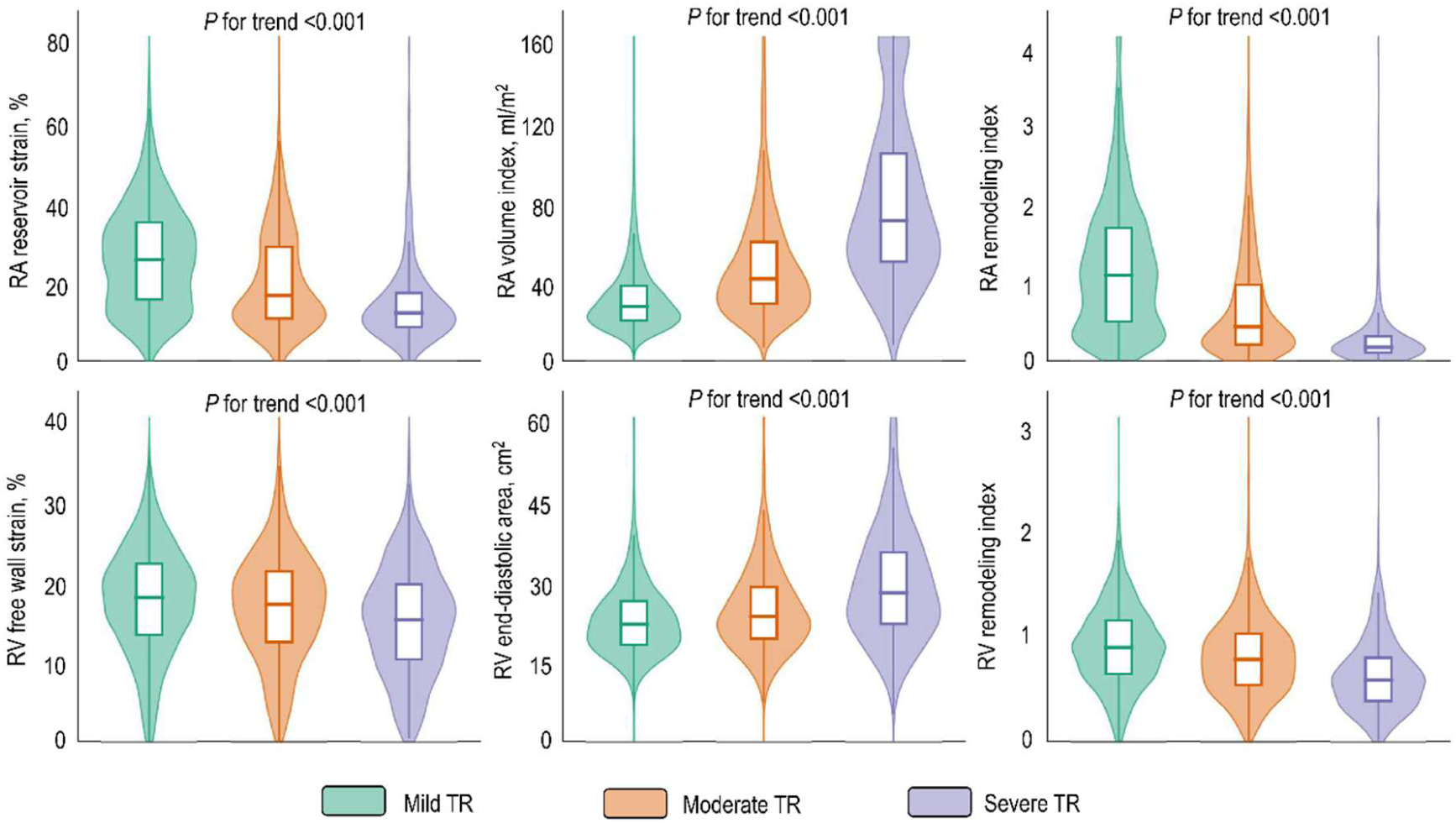
Changes in RA and RV Remodeling Indices Across TR Severity. Violin plots depicting the distributions of RA reservoir strain (RASr), RA volume index (RAVI), RA remodeling index (upper panels), RV free wall strain (RVFWS), RV end-diastolic area (RVEDA), and RV remodeling index (lower panels) across mild, moderate, and severe TR. TR = tricuspid regurgitation; RA = right atrium; RV = right ventricle.

### Discriminative Performance for TR Severity

Discriminative performance for TR severity is summarized in **Table S1**. For severe versus non-severe TR, the RA remodeling index (AUC 0.857) significantly outperformed RASr alone (AUC 0.757; ΔAUC +0.100, *P*<0.05), and the RV remodeling index (AUC 0.710) outperformed RVFWS alone (AUC 0.601; ΔAUC +0.109, *P*<0.05). For moderate-or-greater versus mild TR, both indices likewise outperformed their individual strain components (RA: AUC 0.750, ΔAUC +0.074; RV: AUC 0.619, ΔAUC +0.062; both *P*<0.05). Ordinal c-statistics across three-level severity were 0.750 and 0.622 for the RA and RV remodeling indices, respectively. Center-stratified discriminative performance was consistent (**Figure S3**).

### RA–RV Remodeling Phenotypes

Based on median cutoffs, patients were classified into four RA–RV remodeling phenotypes: Preserved (N=2,967), RA-predominant remodeling (N=1,149), RV-predominant remodeling (N=1,149), and Combined remodeling (N=2,966). Baseline characteristics are summarized in **Table S2** and depicted in **Figure 3**. AF prevalence was markedly higher in the RA-predominant (66.2%) and Combined (67.0%) phenotypes compared with the Preserved (9.7%) and RV-predominant (12.4%; *P*<0.001) phenotypes. Patients with Combined remodeling were more frequently female (61.8%), older (median 74.0 years), with higher LA volume index, E/e’, and more frequent loop diuretic and spironolactone use, reflecting a more advanced HF burden. LV ejection fraction was lowest in the Combined phenotype (57.0%) and highest in the Preserved phenotype (62.7%; *P*<0.001).

**Figure 3.**
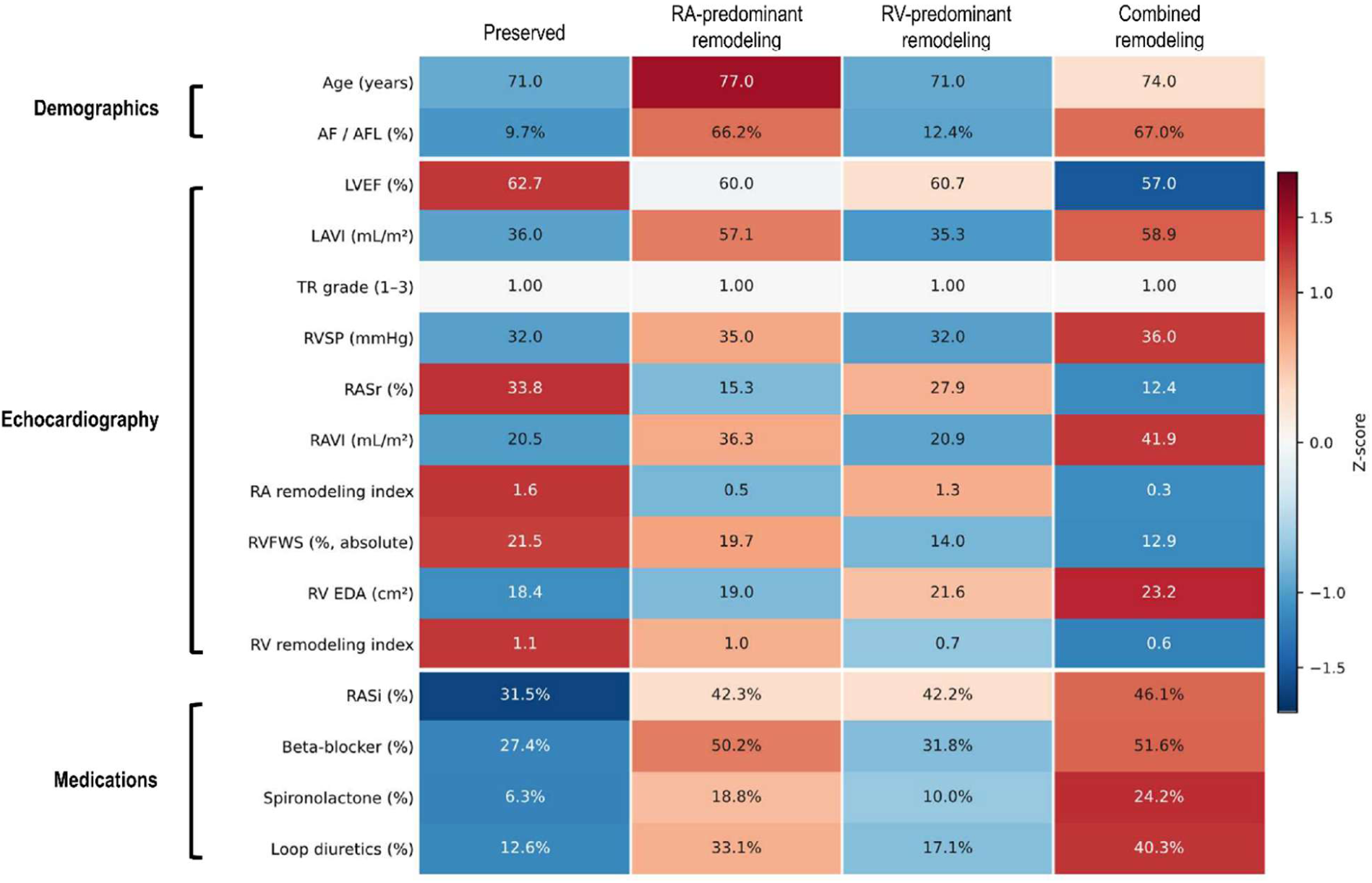
Visual Summary of Baseline Characteristics by RA–RV Remodeling Phenotype. Heatmap summarizing key demographic, echocardiographic, and medication variables across the four RA–RV remodeling phenotypes (Preserved, RA-predominant remodeling, RV-predominant remodeling, and Combined remodeling). Color intensity represents z-score standardized values relative to the overall cohort mean, whereas cell labels indicate observed values, presented as medians for continuous variables and proportions for categorical variables.

### Prognostic Value of RA and RV Remodeling Indices

During follow-up, the composite primary outcome occurred in 574 patients (7.0%), comprising 340 all-cause deaths (4.1%) and 259 HF hospitalizations (3.1%). Cumulative incidence curves are shown in **Figure 4**. Event rates increased progressively across TR severity groups, between low and high index groups (median dichotomized), and across the four phenotypes (all log-rank *P*<0.001). Compared with the Preserved phenotype, adjusted primary outcome risk was higher in the Combined (adjusted HR, 1.79; 95% CI, 1.38–2.33; *P*<0.001) and RV-predominant remodeling phenotypes (adjusted HR, 1.50; 95% CI, 1.12–2.02; *P*=0.007), but did not reach significance in the RA-predominant phenotype (adjusted HR, 1.26; 95% CI, 0.91–1.75; *P*=0.163) (**Table S3**). In univariate Cox regression, both the RA remodeling index (HR per unit decrease, 1.66; 95% CI, 1.47–1.89; *P*<0.001) and RV remodeling index (HR per unit decrease, 4.11; 95% CI, 3.23–5.23; *P*<0.001) were significantly associated with the primary outcome (**Table 2**). After multivariate adjustment, both the RA remodeling index (adjusted HR, 1.27; 95% CI, 1.09–1.49; *P*=0.002) and RV remodeling index (adjusted HR, 2.32; 95% CI, 1.76–3.06; *P*<0.001) retained independent prognostic significance. On mutual adjustment, the RV remodeling index retained independent significance (adjusted HR, 2.21; 95% CI, 1.63–2.98; *P*<0.001), whereas the RA remodeling index was attenuated and no longer significant (adjusted HR, 1.07; 95% CI, 0.91–1.25; *P*=0.431). Among individual components, RASr, RVEDA, and RVFWS were independently associated with the primary outcome, whereas RAVI was not (*P*=0.351; **Table 2**). Results were consistent in competing-risk sensitivity analyses (**Table S4**).

**Figure 4.**
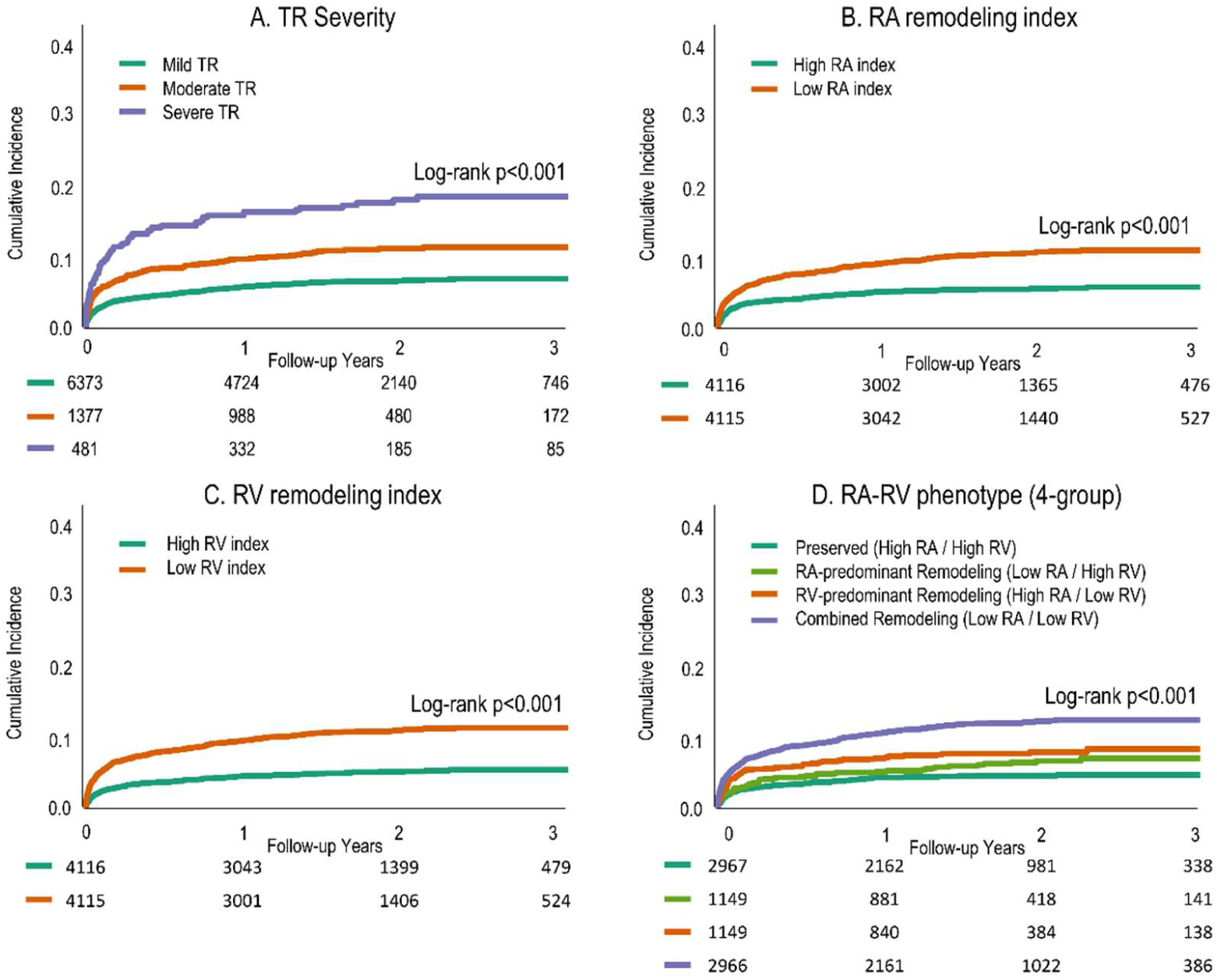
Risk Curves of Primary Outcome According to TR Severity and RA-RV Phenotypes. Kaplan-Meier curves for the composite outcome of all-cause death and HF hospitalization stratified by (A) TR severity, (B) RA remodeling index dichotomized by median cutoff of 0.87, (C) RV remodeling index dichotomized by median cutoff of 0.84, and (D) the four RA–RV remodeling phenotypes defined by these median cutoffs, with the Preserved phenotype as the reference. HF = heart failure; TR = tricuspid regurgitation.

**Table 2.** Association of Individual Echocardiographic Parameters and Remodeling Indices with Clinical Outcomes.

| Variable | Univariate HR (95% CI) | *P* | Multivariate HR* (95% CI) | *P* |
| --- | --- | --- | --- | --- |
| Individual Echocardiographic Parameters |  |  |  |  |
| RA volume index, per 10 mL/m<sup>2</sup> increase | 1.07 (1.05–1.10) | <0.001 | 1.02 (0.98–1.05) | 0.351 |
| RA reservoir strain, per 1% decrease | 1.04 (1.03–1.05) | <0.001 | 1.03 (1.02–1.04) | <0.001 |
| RV end-diastolic area, per 5 cm<sup>2</sup> increase | 1.19 (1.14–1.25) | <0.001 | 1.09 (1.03–1.16) | 0.005 |
| RV free wall strain, per 1% decrease | 1.07 (1.05–1.08) | <0.001 | 1.04 (1.02–1.05) | <0.001 |
| Integrated Remodeling Indices |  |  |  |  |
| RA remodeling index, per unit decrease | 1.66 (1.47–1.89) | <0.001 | 1.27 (1.09–1.49) | 0.002 |
|  |  |  | 1.07 (0.91–1.25)<sup>†</sup> | 0.431 |
| RA remodeling index, per 1-SD decrease | 1.53 (1.38–1.70) | <0.001 | 1.22 (1.07–1.39) | 0.002 |
|  |  |  | 1.06 (0.92–1.21)<sup>†</sup> | 0.431 |
| RV remodeling index, per unit decrease | 4.11 (3.23–5.23) | <0.001 | 2.32 (1.76–3.06) | <0.001 |
|  |  |  | 2.21 (1.63–2.98)<sup>‡</sup> | <0.001 |
| RV remodeling index, per 1-SD decrease | 1.74 (1.58–1.91) | <0.001 | 1.39 (1.25–1.55) | <0.001 |
|  |  |  | 1.36 (1.21–1.53)<sup>‡</sup> | <0.001 |

\*Adjusted for age, sex, atrial fibrillation, LV ejection fraction, TR grade.
<sup>†</sup>Additionally adjusted for RV remodeling index
<sup>‡</sup>Additionally adjusted for RA remodeling index
Abbreviations: CI, confidence interval; HR, hazard ratio; RA, right atrium; RV, right ventricle; SD, standard deviation; TR, tricuspid regurgitation.

For secondary outcomes, only the RV remodeling index was independently associated with all-cause death (adjusted HR, 1.85; 95% CI, 1.31–2.62; *P*<0.001; **Table S5**). Both indices were independently associated with HF hospitalization (RA remodeling index: adjusted HR, 3.76; 95% CI, 2.61–5.43; *P*<0.001; RV remodeling index: adjusted HR, 3.24; 95% CI, 2.08–5.03; *P*<0.001), with consistent associations across competing-risk specifications incorporating all-cause death and TV surgery as competing events.

### Incremental Prognostic Value

Incremental prognostic performance beyond the reference clinical model (age, sex, AF, LV ejection fraction, TR grade; c-statistic 0.710, AUC 0.713) is detailed in **Table 3** and **Table S6**. Adding the RV remodeling index alone (Model 4) significantly improved discrimination across all metrics (Δc-statistic +0.010, *P*=0.017; ΔAUC +0.013, *P*=0.004; NRI +0.237, *P*<0.001). Adding both RA and RV remodeling indices (Model 5) yielded similar incremental gains (Δc-statistic +0.010, *P*=0.019; ΔAUC +0.014, *P*=0.004; NRI +0.214, *P*<0.001). The combination of RASr and RVFWS alone provided the most pronounced incremental discrimination (Δc-statistic +0.013, *P*=0.005; ΔAUC +0.016, *P*=0.001; NRI +0.295, *P*<0.001), whereas RAVI alone contributed no significant incremental discrimination across any metric.

**Table 3.**
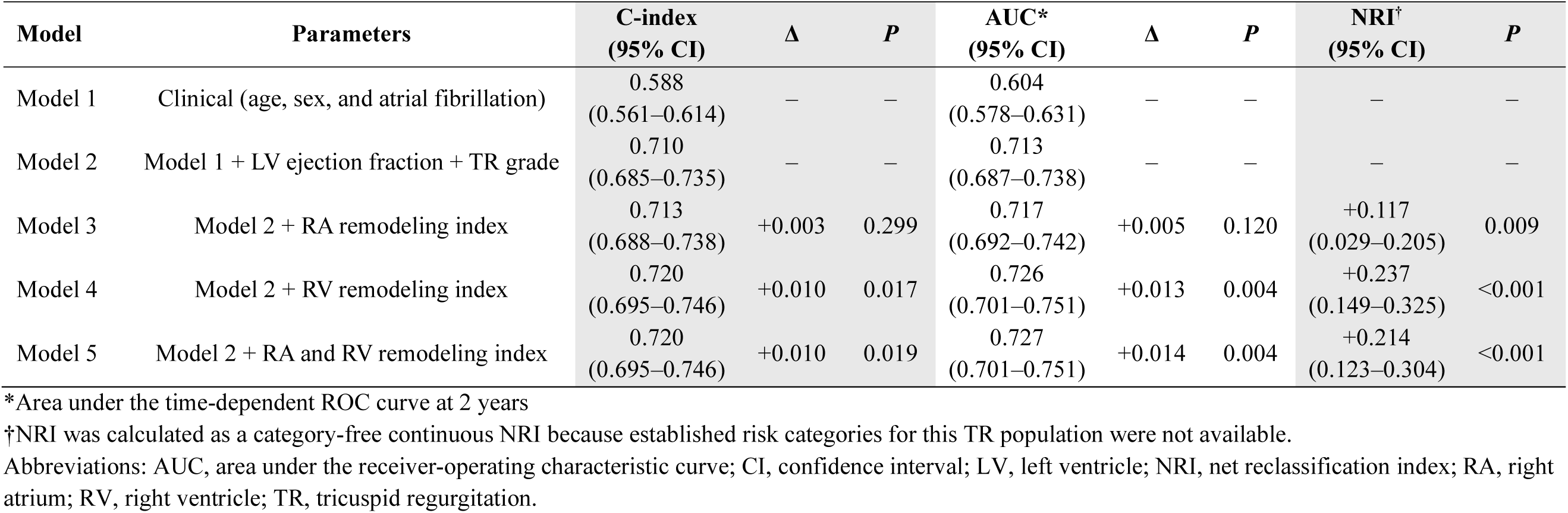
Incremental Prognostic Value of RA and RV Remodeling Indices.

| Model | Parameters | C-index<br>(95% CI) | $\Delta$ | *P* | AUC*<br>(95% CI) | $\Delta$ | *P* | NRI†<br>(95% CI) | *P* |
| --- | --- | --- | --- | --- | --- | --- | --- | --- | --- |
| Model 1 | Clinical (age, sex, and atrial fibrillation) | 0.588<br>(0.561–0.614) | – | – | 0.604<br>(0.578–0.631) | – | – | – | – |
| Model 2 | Model 1 + LV ejection fraction + TR grade | 0.710<br>(0.685–0.735) | – | – | 0.713<br>(0.687–0.738) | – | – | – | – |
| Model 3 | Model 2 + RA remodeling index | 0.713<br>(0.688–0.738) | +0.003 | 0.299 | 0.717<br>(0.692–0.742) | +0.005 | 0.120 | +0.117<br>(0.029–0.205) | 0.009 |
| Model 4 | Model 2 + RV remodeling index | 0.720<br>(0.695–0.746) | +0.010 | 0.017 | 0.726<br>(0.701–0.751) | +0.013 | 0.004 | +0.237<br>(0.149–0.325) | <0.001 |
| Model 5 | Model 2 + RA and RV remodeling index | 0.720<br>(0.695–0.746) | +0.010 | 0.019 | 0.727<br>(0.701–0.751) | +0.014 | 0.004 | +0.214<br>(0.123–0.304) | <0.001 |

\*Area under the time-dependent ROC curve at 2 years
†NRI was calculated as a category-free continuous NRI because established risk categories for this TR population were not available.
Abbreviations: AUC, area under the receiver-operating characteristic curve; CI, confidence interval; LV, left ventricle; NRI, net reclassification index; RA, right atrium; RV, right ventricle; TR, tricuspid regurgitation.

### Subgroup Analyses

Pre-specified subgroup analyses were performed by TR severity (mild vs. moderate-or-greater), AF status, pulmonary hypertension status (RVSP <40 vs. ≥40 mmHg), and diuretic use (**Figure 5**). The RA remodeling index showed a significant TR severity interaction (interaction *P*=0.043), with stronger association in moderate-or-greater TR (adjusted HR 2.04; 95% CI, 1.42–2.95) than in mild TR (1.13; 0.95–1.34). It was also significantly associated with outcomes in patients with sinus rhythm (adjusted HR 1.26; 95% CI, 1.06–1.49) and on diuretics (1.33; 1.05–1.69), without significant interaction by AF status or diuretic use. The RV remodeling index showed a similar TR severity interaction (interaction *P*=0.011), with markedly stronger association in moderate-or-greater TR (adjusted HR 5.09; 95% CI, 3.15–8.24) than in mild TR (1.64; 1.17–2.30), and consistent prognostic value regardless of AF status, pulmonary hypertension, and diuretic use.

**Figure 5.**
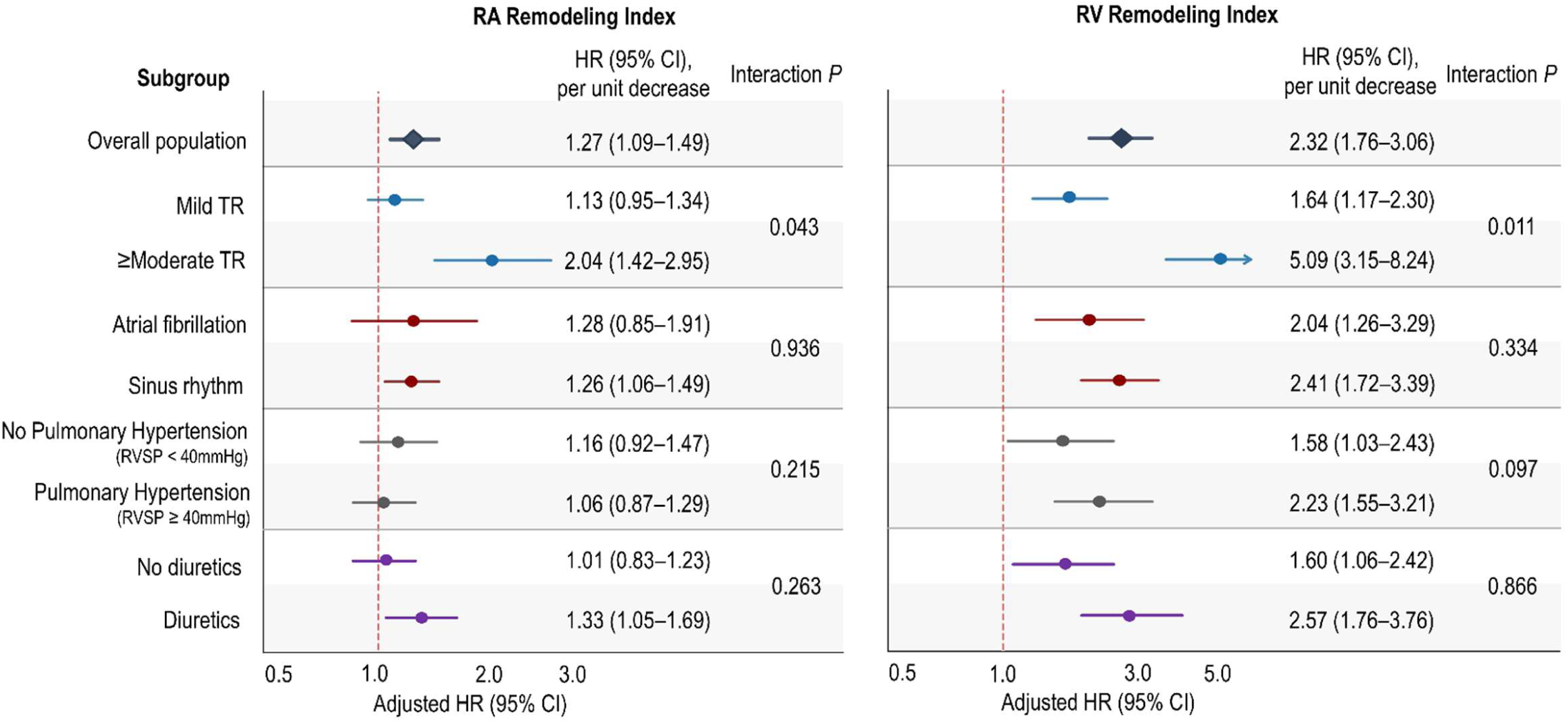
Subgroup Analysis for RA and RV Remodeling Indices. Forest plots depicting adjusted hazard ratios (HRs) with 95% confidence intervals (CIs) for the RA remodeling index (left panel) and RV remodeling index (right panel) per unit decrease across pre-specified subgroups. All models were adjusted for age, sex, atrial fibrillation, LV ejection fraction, and TR grade. RVSP = RV systolic pressure; LV = left ventricle; HR = hazard ratio; CI = confidence interval.

To further examine the prognostic gradient of each index, we evaluated dichotomized index segments (**Figure S4**). Prognostic significance of both indices was pronounced in the low-index segments (RA remodeling index: adjusted HR, 2.68; 95% CI, 1.54–4.64; *P*<0.001; RV remodeling index: adjusted HR, 2.98; 95% CI, 1.80–4.95; *P*<0.001). An apparent increase in all-cause mortality observed in 43 patients in the high RA and high RV index segment at SNUH was largely non-cardiovascular in etiology (40/43 [93%] non-cardiovascular deaths, predominantly malignancy, sepsis, and respiratory failure; **Table S7**), and was interpreted as reflecting sparse non-cardiac events rather than a biological inverse association. Both indices remained independent prognostic predictors in center-adjusted sensitivity analysis (RA remodeling index: adjusted HR, 1.21; 95% CI, 1.04–1.42; *P*=0.015; RV remodeling index: adjusted HR, 2.28; 95% CI, 1.72–3.01; *P*<0.001) and center-stratified analysis (RA remodeling index: adjusted HR, 1.21; 95% CI, 1.03–1.41; *P*=0.016; RV remodeling index: adjusted HR, 2.27; 95% CI, 1.72–3.00; *P*<0.001) (**Table S8**).

## DISCUSSION

In this multicenter study of 8,231 patients with functional TR, we developed integrated RA and RV remodeling indices combining strain-derived function with structural measures, derived through fully automated AI-based echocardiographic analysis. The indices outperformed individual strain parameters for TR severity discrimination. Although both were independently associated with the primary outcome after clinical adjustment, the RV index emerged as the dominant prognostic marker on mutual adjustment and provided incremental value beyond a standard clinical model. The RA index was most informative for atrial-stage characterization and demonstrated prognostic relevance in selected clinical contexts, supporting complementary roles of the two indices in TR assessment.

### RA Remodeling Index: TR Severity Discrimination and Stage Biology

The RA remodeling index (RASr/RAVI) outperformed RASr alone for severe TR discrimination (AUC 0.857 vs. 0.757), likely reflecting the complementary nature of its components: RAVI captures the structural burden of RA volume overload, whereas RASr reflects atrial myocardial functional reserve. Because RA dilation often precedes functional impairment, with myocardial dysfunction predominating later, the integrated index may capture stage-dependent remodeling more sensitively than either parameter alone. This aligns with the pathophysiologic staging model of atrial secondary TR by Muraru et al.(22)

Clement et al. defined the RA stiffness index as the mathematical inverse of our RA remodeling index (RAVI/RASr); like our index, it lost independent significance after multivariable adjustment incorporating RV function.(20) This convergence suggests that the independent prognostic contribution of RA functional reserve is most evident in specific clinical contexts, notably moderate-or-greater TR and sinus rhythm, as supported by our interaction analyses.

Low RA remodeling index values predominated in phenotypes with high AF prevalence, consistent with AF-induced atrial myopathy impairing RASr while promoting RA dilation, thereby lowering the index independent of TR severity.(11,12) The significant interaction with TR severity (interaction *P*=0.043), with a stronger association in moderate-or-greater TR, supports the interpretation of the RA remodeling index as a disease-stage marker rather than a simple severity surrogate.(5,22) Accordingly, the index may be best characterized as a marker of atrial-stage remodeling and disease burden, with prognostic relevance most pronounced in HF hospitalization and selected clinical contexts (moderate-or-greater TR, sinus rhythm).

### RV Remodeling Index: Independent Prognostic Value

The RV remodeling index (RVFWS/RVEDA) provided significant incremental prognostic value beyond the reference clinical model and was the dominant predictor on mutual adjustment. Prior studies established RVFWS as an independent prognostic marker in TR,(9,10) and ratio-based approaches such as the RVFWS-to-pulmonary pressure ratio have been proposed as load-corrected functional measures.(21) Unlike strain-to-pressure ratios, the RV remodeling index normalizes functional impairment to structural chamber remodeling, a distinction particularly relevant in TR where volume overload drives progressive chamber enlargement even with relatively preserved systolic function. This indexing preserves the prognostic signal of longitudinal RV dysfunction while accounting for structural burden.

The markedly stronger association in moderate-or-greater TR (HR 5.09 vs. 1.64 in mild TR; interaction *P*=0.011) suggests that RV functional reserve becomes critical once structural remodeling is established, and that the integrated index captures this threshold effect more effectively than RVFWS alone. Unlike the RAVC_STE index (RASr/RVFWS), which Clement et al. reported to underperform volumetric coupling indices in secondary TR,(20) our RV remodeling index normalizes RVFWS to RV chamber area rather than to RA strain, suggesting that correcting for structural RV remodeling better captures the prognostically relevant dimension of RV dysfunction.

The RV remodeling index showed consistent prognostic value across AF status, pulmonary hypertension, and diuretic use, underscoring its robustness across heterogeneous TR presentations. The RV index retained independent significance for all-cause death (adjusted HR 1.85; *P*<0.001), whereas the RA index did not; conversely, the RA index was strongly associated with HF hospitalization (adjusted HR 3.76; *P*<0.001). This divergence likely reflects distinct hemodynamic roles, with RV dysfunction conferring broader mortality risk and atrial remodeling specifically predisposing to HF hospitalization.

### RA–RV Remodeling Phenotypes: Clinical Characterization and Implications

The four phenotypes showed clinically distinct profiles with progressively divergent outcome risk. The RA-predominant and Combined phenotypes had markedly higher AF prevalence (66.2% and 67.0%), consistent with AF-induced atrial myopathy as the primary driver of RA remodeling in the absence of severe structural TR.(5,22) The RV-predominant phenotype showed relatively preserved RA function but higher RVSP, suggesting pressure overload as the dominant driver of RV remodeling. The Combined phenotype was characterized by older age, lower LV ejection fraction, higher filling pressures, and substantially greater diuretic burden, reflecting the most advanced RA and RV decompensation.

This framework aligns conceptually with the established atrial-versus-ventricular secondary TR classification,(22) where the RA-predominant phenotype corresponds to atrial secondary TR (with AF but preserved RV function) and the RV-predominant and Combined phenotypes reflect the spectrum of ventricular secondary TR with progressive RV dysfunction. However, our classification extends beyond etiologic dichotomization by quantifying the relative degree of atrial and ventricular functional impairment, enabling graded risk stratification across four clinically distinct profiles and providing clinically actionable information beyond TR severity grading alone.

### Role of AI-Based Automated Measurement

A key enabling feature was the use of a fully automated AI-based platform for derivation of all RA and RV parameters without any manual interaction. Manual measurement of RASr and RVFWS is time-consuming, operator-dependent, and subject to substantial inter-observer variability, limiting systematic application in routine practice. The platform used here has been previously validated across cardiovascular conditions,(13–18) and achieved 96.2% feasibility in this real-world multicenter cohort. Automated AI-derived quantification is a prerequisite for broader clinical adoption of integrative right-heart indices in TR management;(23) this study demonstrates the feasibility of automated derivation of integrated RA and RV remodeling indices at scale.

### Limitations

This study has several limitations. First, the retrospective design from two tertiary academic centers may limit generalizability to broader populations and community-based settings. Second, TR severity grading was based on integrated clinical judgment, reflecting real-world practice but potentially introducing classification variability. Third, the median cutoff values for phenotype classification were derived from the study cohort distribution and require external validation. Fourth, the relatively short median follow-up may underestimate long-term outcome differences, particularly for lower-risk phenotypes. Fifth, all-cause mortality may include non-cardiovascular deaths unrelated to right-heart remodeling, potentially attenuating the biological association between indices and cardiovascular risk. Finally, the prognostic value of these indices in the context of TV intervention, and their potential utility as treatment targets or response markers, remains to be established.

### Conclusions

In this large multicenter cohort of patients with functional TR, automated AI-derived integrated RA and RV remodeling indices improved TR severity discrimination and enabled clinically meaningful right-heart phenotyping. The RA remodeling index was most informative for atrial-stage characterization and disease burden, whereas the RV remodeling index served as the principal prognostic marker with incremental value beyond conventional clinical parameters. Four phenotypes based on these indices showed progressively divergent outcome risk. Together, these findings support a comprehensive integrated right-heart remodeling framework and highlight the complementary roles of RA and RV remodeling in TR characterization and risk stratification.

## Data Availability

The data underlying this study cannot be made publicly available due to ethical restrictions set by the IRB of the study institution; i.e., public availability would compromise patient confidentiality and participant privacy. Please contact the corresponding author to request the minimal anonymized dataset. Researchers with additional inquiries about the deep learning model developed in this study are also encouraged to reach out to the corresponding author.

## Contributors

Jiesuck Park: Conceptualization; Data curation; Formal analysis; Methodology; Visualization; Writing – original draft; Writing – review & editing.

Soongu Kwak: Conceptualization; Data curation; Investigation; Writing – original draft; Writing – review & editing.

Yeonyee E. Yoon: Conceptualization; Funding acquisition; Project administration; Supervision; Writing – review & editing.

Jun-Bean Park: Conceptualization; Project administration; Resources; Supervision; Writing – review & editing.

Jiyeon Kim: Software; Data curation; Methodology.

Jaeik Jeon: Software; Data curation; Methodology. Yeonggul Jang: Software; Data curation; Methodology.

Seung-Ah Lee: Software; Data curation; Methodology.

MinJung Bak: Validation; Investigation; Writing – review & editing.

Hong-Mi Choi: Validation; Investigation; Writing – review & editing.

In-Chang Hwang: Validation; Investigation; Writing – review & editing.

Seung-Pyo Lee: Validation; Investigation; Writing – review & editing.

Hyung-Kwan Kim: Validation; Investigation; Writing – review & editing.

Yong-Jin Kim: Validation; Investigation; Writing – review & editing.

Goo-Yeong Cho: Validation; Investigation; Writing – review & editing.

## Declaration of Interests

Y.E.Y, J.K., J.J., Y.J., and S.A.L. are currently affiliated with Ontact Health Co., Ltd. Y.E.Y holds equity in Ontact Health Co., Ltd. The other authors have no conflicts of interest to declare.

## Acknowledgements

This work was supported by a grant from the Korea government (Ministry of Science and ICT) (No. 2022000972, Development of a Flexible Mobile Healthcare Software Platform Using 5G MEC).

## PERSPECTIVES

### COMPETENCY IN MEDICAL KNOWLEDGE

Automated integration of right atrial and right ventricular strain with chamber geometry may improve characterization of right-heart remodeling in functional tricuspid regurgitation beyond valve-centric severity grading.

### TRANSLATIONAL OUTLOOK

Prospective validation is needed to determine whether RA–RV remodeling phenotypes can guide follow-up intensity, timing of intervention, and response assessment after tricuspid valve therapy.

## ABBREVIATIONS

AF: atrial fibrillation
AUC: area under the receiver-operating characteristic curve
HF: heart failure
RA: right atrial/atrium
RASr: right atrial reservoir strain
RAVI: right atrial volume index
RV: right ventricular/ventricle
RVEDA: right ventricular end-diastolic area
RVFWS: right ventricular free wall strain
TR: tricuspid regurgitation

## Notes

### Author Declarations

The study protocol was approved by the Institutional Review Board of each participating institution (SNUH H-2506-143-1652 and SNUBH B-2605-1047-405), with a waiver of informed consent granted due to the retrospective study design.

